## Supplemental Materials for "Antigen concentration, viral load, and test performance for SARS-CoV-2 in multiple specimen types"

**Supplementary Material**

**Supplementary Material A. Summary of SARS-CoV-2 cases, associated specimens, and test results.**

Result totals are shown for the study participants for index cases, household contacts, and non-household contacts. The proportion of samples selected and run for antigen testing from each participant is shown for nasopharyngeal swab (NPS), anterior nares swab (ANS), and saliva. The lysis buffer for the STANDARD Q ANS specimen was used for antigen concentration determination in the ANS specimen. Because household contacts had multiple timepoints per individual, results reflect the combined individual and timepoint together for the 64 household contacts (total of 224 combinations). Positive and negative classifications correspond to available test results from laboratory PCR results, SalivaDirect, STANDARD Q Saliva, LumiraDx, and STANDARD Q point-of-care (anterior nasal) and exclude antigen concentration measurement results.

| **Participant type** | **Negative by all tests** | **Positive by only one test** | **Positive by only two tests** | **Positive by three or more tests** | **Sample type** | **Selected for antigen concentration testing** | **Insufficient or missing sample** | **Antigen concentration determined** |
| --- | --- | --- | --- | --- | --- | --- | --- | --- |
| **Close Contacts,**  **Household**  **224 individuals and timepoint combinations from 64 individuals)** | 143 | 19 | 15 | 47 | **NPS** | 191 | 28  (no NPS taken) | 163 |
|  |  |  |  |  | **ANS** | 191 | 5 | 186 |
|  |  |  |  |  | **Saliva** | 191 | 8 | 183 |
| **Close Contacts, Non-household**  **(150)** | 117 | 6 | 8 | 19 | **NPS** | 94 | 0 | 94 |
|  |  |  |  |  | **ANS** | 94 | 1 | 93 |
|  |  |  |  |  | **Saliva** | 94 | 2 | 92 + 2 additional not selected |

**Supplementary Material B. Case definitions for the study.**

*Symptomatic*: Participants presenting with cough, shortness of breath, difficulty breathing, or at least two of the following symptoms at the time of sampling: fever, chills, rigor, myalgia, headache, sore throat, new olfactory or taste disorder.

*Oligosymptomatic:* Participants who presented with one or more mild symptoms but did not fit the symptomatic case definition and reported no care seeking or changes to behavior.

*Asymptomatic:* Participants with no symptoms at the time of sampling.

**Supplementary Material C. Score card for line intensity on the STANDARD Q COVID-19 Ag test (0 refers to no visible test line, or negative score) for nasal and saliva.**

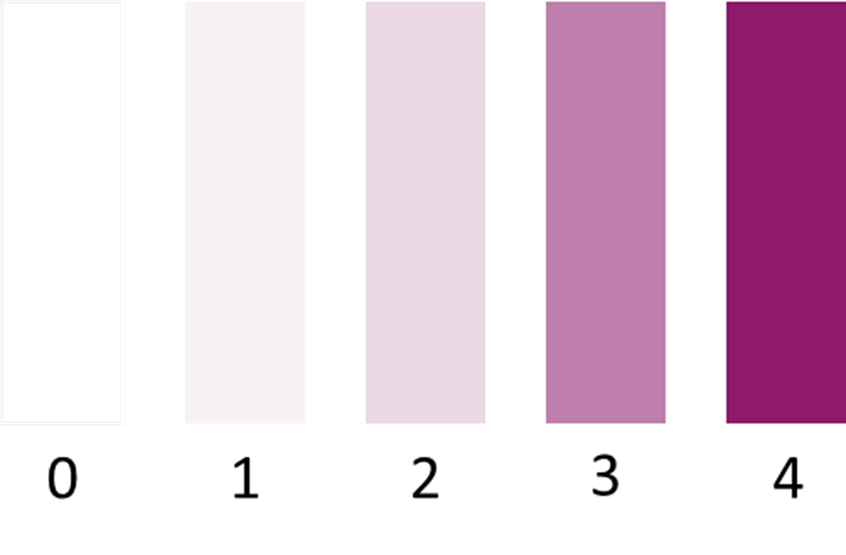

**Supplementary Material D**. Specimens for which there was a high viral load in the NPS but a negative test result from the antigen test conducted on the ANS specimen. For binary test results, 1 = positive, 0 = negative. Test line intensity for STANDARD Q Nasal and Saliva tests were reported according to scale shown in Supplementary Material C (0-4). Lumira Signal refers to signal output from the LumiraDx instrument.

| Participant ID | Visit # | PCR N gene, Ct | Viral genome copies/mL | Sequencing clade | | N-antigen sense mutations | STANDARD Q Saliva Result | STANDARD Q Saliva Intensity | Lumira Result | Lumira Signal | STANDARD Q Nasal Result | STANDARD Q Nasal Intensity | Antigen Concentration, pg/mL in Standard Q buffer |
| --- | --- | --- | --- | --- | --- | --- | --- | --- | --- | --- | --- | --- | --- |
| CC-088 | 1 | 22.2 | 30188058 | Gamma, V3 | | P80R, R203K, G204R | 1 | 4 | 1 | 1181.6 | 0 | 0 | 29.2 |
| CC-209 | 1 | 23.5 | 2627412 | not available | | not available | 0 | 0 | 0 | 252.4 | 0 | 0 | 0.0 |
| CC-135 | 1 | 23.7 | 10720679 | Gamma, V3 | | P80R, R203K, G204R, T391I | 1 | 1 | 0 | 518.5 | 1 | 1 | 20.6 |
| CC-017 | 3 | 23.8 | 10225694 | Gamma, V3 | | P80R, R203K, G204R, Q240L | 0 | 0 | 0 | 245.9 | 0 | 0 | 0.0 |
| CC-163 | 1 | 25.4 | 3267859 | Gamma, V3 | | P80R, R203K, G204R, P344L | 0 | 0 | 0 | 555.1 | 1 | 1 | 4.2 |
| CC-114 | 1 | 25.9 | 2319762 | Gamma, V3 | | P80R, R203K, G204Q | 0 | 0 | 0 | 321.6 | 1 | 1 | 258.9 |
| CC-043 | 1 | 25.7 | 2593367 | Gamma, V3 | | P80R, R203K, G204R | 1 | 3 | 0 | 293.5 | 0 | 0 | 0.0 |
| CC-054 | 1 | 25.9 | 2341293 | Gamma, V3 | | P80R, T135I, R203K, G204R | 1 | 1 | 0 | 780.1 | 1 | 1 | 84.8 |
| IND-037 | 1 | 27.1 | 1017678 | Gamma, V3 | P80R, R203K, G204R, S413I | | 0 | 0 | 0 | 661.4 | 1 | 1 | 39.9 |
| CC-009 | 2 | 29.5 | 717496 | not available | not available | | 0 | 0 | 1 | 1690.2 | 0 | 0 | 79.1 |
| CC-200 | 1 | 29.6 | 158489 | not available | not available | | 0 | 0 | 0 | 255.9 | 0 | 0 | 0.0 |
| CC-029 | 2 | 34.3 | 16670 | not available | not available | | 1 | 2 | 0 | 243.4 | 0 | 0 | 0.0 |
| CC-082 | 1 | 33.6 | 10755 | not available | not available | | 0 | 0 | 0 | 185.7 | 0 | 0 | 7.6 |
| CC-149 | 1 | 33.5 | 11497 | not available | not available | | 0 | 0 | 0 | 232.2 | 0 | 0 | 0.0 |
